# SenseCheQ: Home-based Nerve Function Self-Assessment using Autonomous Quantitative Sensory Testing

**DOI:** 10.64898/2026.04.15.26350779

**Authors:** Johannes Gausden, Marin Dujmović, James P Dunham, Bhushan Thakkar, Thomas Bennett, Charlotte Burgess, Mathew Barraclough, Alan Young, Roger G Whittaker, Timothy Robinson, Lesley Colvin, Anthony O’Neill, Anthony E Pickering

## Abstract

Neuropathy caused by chemotherapy is a common and debilitating side-effect of cancer treatment. Approximately 30% of patients experience chronic neuropathy and the lack of effective treatments means that early detection of neuropathy to trigger chemotherapy regime modification remains the best option for prevention. Early detection is challenging because no diagnostic tools exist with sufficient precision and accessibility for patient/clinical adoption. To tackle this problem, we developed SenseCheQ to enable self-administered autonomous sensory testing which can be used by patients at home. We present the engineering approach taken to address this challenge, including novel thermo-haptic integration allowing skin temperature clamping and precise haptic stimulus calibration to improve reproducibility. Robust performance is demonstrated in the context of environmental and user-related variability. Additionally, we share prospective case studies of people undergoing chemotherapy treatment for cancer, conducting regular sensory testing with SenseCheQ to monitor their nerve function at home. These proof-of-principle studies show SenseCheQ can detect changes in nerve function, matching patient reported outcomes. This illustrates the promise of SenseCheQ as a scalable platform for neuropathy-detection in the community.

## Introduction

Chemotherapy-induced peripheral neuropathy (CIPN) is a common and debilitating adverse side-effect of many systemic anti-cancer agents, including taxanes^1,2^, platin compounds^3,4^, vinca alkaloids^5,6^ and bortezomib^7,8^, which impact Aβ, Aδ and C fibre sensory function. Up to 70% of patients develop acute neuropathy during treatment (including pain, numbness, fine motor impairment and autonomic dysfunction (constipation, incontinence and postural hypotension) frequently leading to chemotherapy dose reductions, treatment delays, or treatment discontinuation altogether^9^. In 30% of patients the symptoms of neuropathy last for over 6 months and the damage can be life-long^9,10^. Despite its high prevalence and the long term impact on quality-of-life impairing cancer survivorship, there are no proven preventive or efficacious remedial therapies for CIPN beyond modifying or ceasing chemotherapy^10,11^. Early diagnosis of CIPN is therefore important, allowing timely chemotherapy regime modification to prevent further neurotoxicity and maximise the potential for recovery. However, early diagnosis is hampered by the fact that current clinical assessments rely heavily on patient report, and that bedside clinical tests of neural function lack sensitivity^12–14^.

Quantitative sensory testing (QST) offers a more objective psychophysical assessment of nerve function^15–18^ and has been shown to be able to detect changes due to neurotoxic chemotherapy^19,20^. However, despite these promising results, QST has not been adopted in routine clinical oncology practice. There are two main reasons for this lack of adoption. Firstly, practical issues, which include the need for trained personnel, time-intensive testing protocols, relatively high equipment costs, and the requirement for patient attendance at a clinical/lab facility at a particularly challenging time in their cancer treatment journey. The second obstacle to adoption is the lack of well-powered studies to provide compelling evidence of utility of QST for early detection of CIPN with adequate sensitivity and specificity^21^. Such studies would require regular longitudinal testing during chemotherapy treatment (ideally at least once per treatment cycle) which is challenging given the practical obstacles noted above. Simplified “bedside” QST protocols address equipment expense but still require the presence of a skilled practitioner and lack the sensitivity required to be suitable for early detection of nerve damage^13,14,22–24^.

To tackle these obstacles, QST needs to made easy to deploy, relatively quick to administer and cost-effective. Additionally, it should perform reliably across a wide range of environmental conditions as encountered in home settings. Is should also ideally allow self-administered testing by patients, shifting the locus of control, supporting autonomy, and reducing burden.

Working with patient partners, we iteratively defined such a protocol, informing the development of SenseCheQ: an integrated stimulation and assessment platform, designed to deliver reliable sensory testing of thermal and mechanical sensitivity in home environments. Through haptic-stimulation calibration, innovative stimulator integration, adaptive algorithms and patient-centred interface design, SenseCheQ minimizes environmental and procedural variability while maintaining sensitivity to physiological and pathophysiological changes. In this paper we describe the technological approaches underpinning SenseCheQ, as well as results from a home study designed to measure reliability and feasibility in healthy adults. Finally, we present two exemplar case studies of people using SenseCheQ whilst undergoing chemotherapy for cancer; assessing their own sensory function unsupervised at home during their treatment cycles.

## Results

### The SenseCheQ Specification

The design brief for SenseCheQ was informed by the standardised QST protocol developed by the German Research Network on Neuropathic Pain (DFNS)^18^. Our own preliminary studies^20^, alongside discussions and feedback from the SenseCheQ Patient Partner group resulted in a modified DFNS protocol designed to deliver a concise but potentially informative set of tests. The reduced test set of vibration, cold and warm detection thresholds was defined to identify changes in Aβ, Aδ, and C nerve fibre function, respectively^19^, given that different types of chemotherapy can produce predominantly large or small fibre dysfunction. Again, informed by input from patient partners, we investigated the suitability of various test locations^20^. The thenar eminence was found to be a better test site than the more commonly targeted dorsum or sole of the foot due to higher sensitivity to thermal and mechanical stimulation, less age-related change, and better accessibility. This decision was corroborated by the observation of larger detection-threshold differences at the thenar eminence than the dorsum of the foot when comparing between people with CIPN and age matched controls^20^.

Building on this framework, the hardware and software of SenseCheQ were designed to create an automated test protocol for home use. The design was created iteratively with input and feedback from patient partners to ensure clarity and intuitive use. For example, most of the patient partners were against the use of a smartphone, app-based interface with Bluetooth connectivity for reasons of inclusivity and favoured physical buttons rather than a touchscreen interface. This aligns with the evidence showing that proficiency with touchscreen interfaces declines with age, fatigue, motor and neurological impairment^25–28^, particularly relevant issues for people with cancer who are undergoing arduous chemotherapy treatments and who are typically in an older age range. Accordingly, we designed the interface such that users interact with SenseCheQ via two haptic push buttons (Fig 1A and B). The left (green) button is used to progress through the test program (i.e. initialize testing) whilst the right (red) button is used to respond as soon as a change in sensory percept is detected (start of vibration, cooling or warming). This physical interface ensures lower latency and reduced variability in response-time-sensitive tasks^29,30^. During each test session, the user interface guides users through four measurements of the three parameters: vibration, cold and warm detection thresholds (VDT, CDT, and WDT, respectively) (Fig 1C). This simplified design and protocol improves device accessibility for those less comfortable with digital technologies. Participants in the home-based testing trial commented on the ease of use, intuitive nature of the interface and simplicity of home testing (Fig 1D).

**Fig 1.**
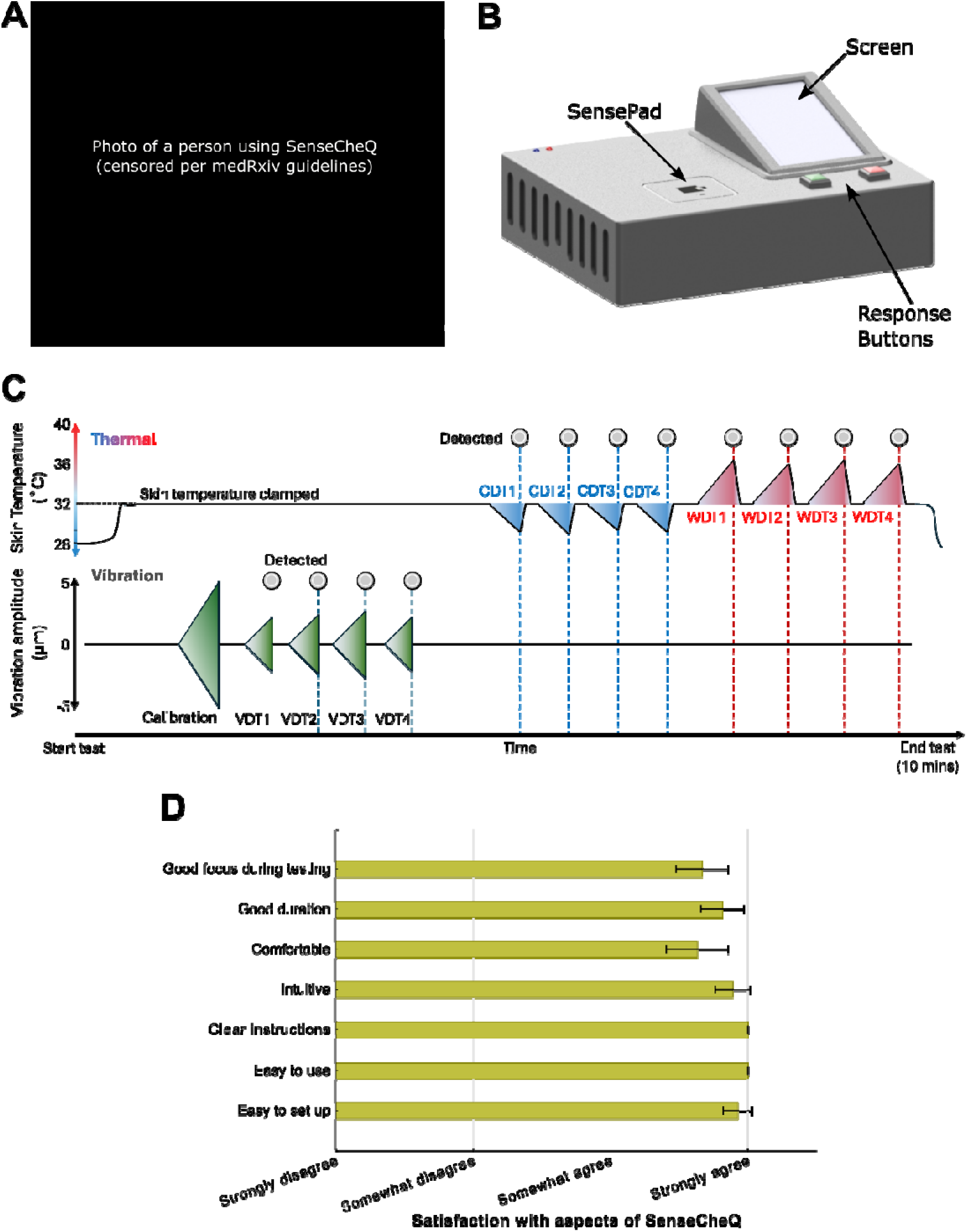
SenseCheQ, protocol and user feedback. A – SenseCheQ being used by a study participant, note the left hand (thenar eminence) located over the SensePad. B – rendered image of the SenseCheQ device. C – Schematic of the test protocol. Skin temperature at the thenar eminence is stabilized to 32°C which is followed by the haptic calibration routine. Users complete four measurements within the session of vibration, warm and cool detection thresholds evoked in response to ramping stimuli. D – Participant (N = 20) feedback on ease-of-use of SenseCheQ (bars represent means and spreads indicate 95% CI).

Careful consideration was also given to identify sources of variability which could be engineered out of the measurements. To this end, a first design innovation came from the decision to create an integrated “*SensePad*” capable of delivering both mechanical and thermal stimulation to the thenar eminence. The *SensePad* (Fig 1B) is the top surface of the hardware *Stack* assembly (Fig 2B) which can vibrate due to the linear resonant actuator (LRA) located at the bottom. Thermal control and stimulation are provided by a Peltier module (12.3 mm x 12.3 mm), which is mounted underneath an Aluminium thermal faceplate. Two Pt1000 thermometers and a 3-axis accelerometer mounted within the stack-top provide feedback for proportional-integral (PI) thermal control, thermal safety hardware and a haptic calibration routine (detailed later), respectively.

**Fig 2.**
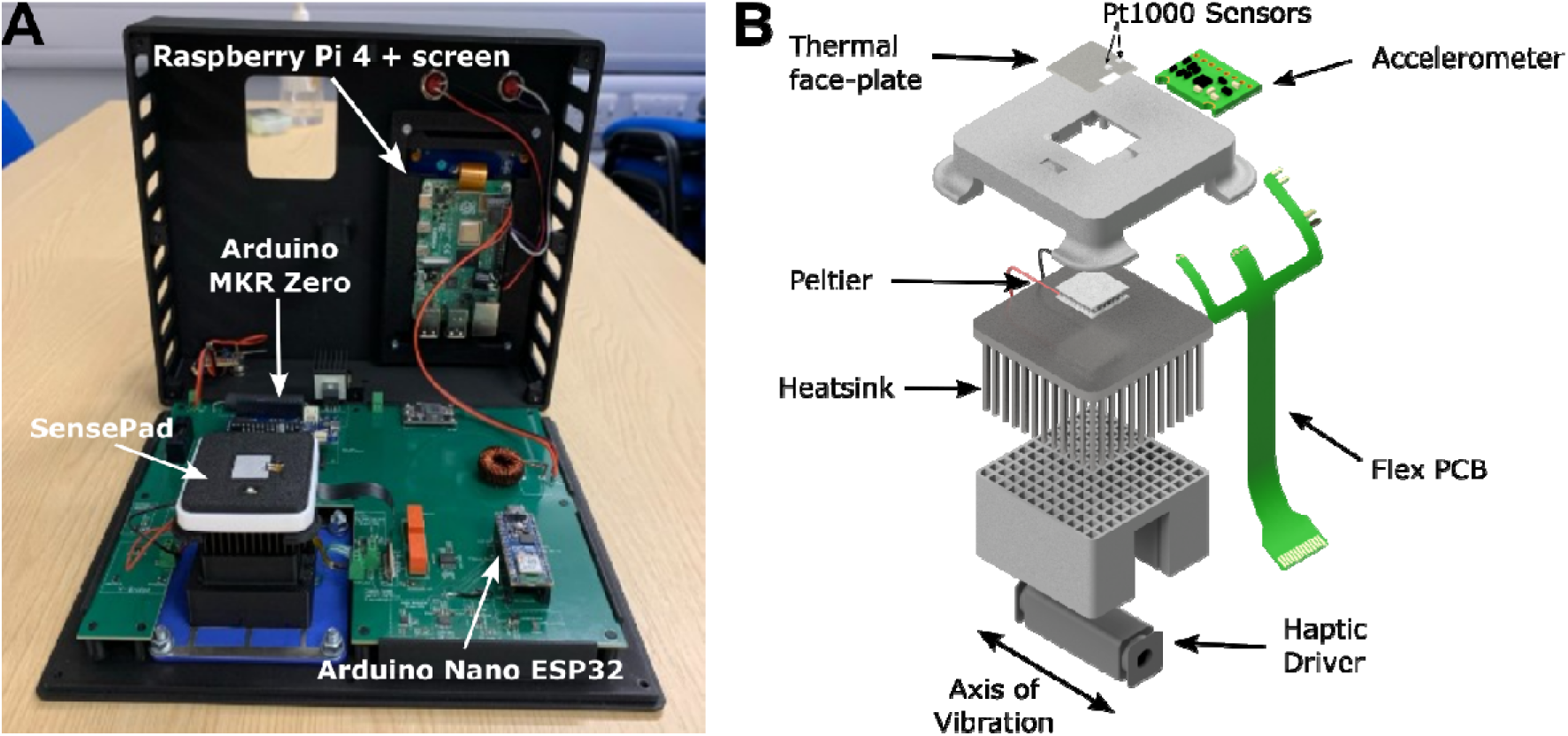
SenseCheQ internals and exploded view of the SensePad stack. A – Internal layout of SenseCheq with the SensePad, Arduino Nano ESP 32 (vibration stimulation microcontroller), Arduino MKR Zero (thermal stimulation microcontroller) and Raspberry Pi (UI and main control of session flow) highlighted. B – The top of the stack houses the vibration-feedback and the thermal hardware. Vibration stimulation is supplied by a haptic actuator, which oscillates laterally and is housed within the bracket at the bottom of the stack.

A third Pt1000 measures local skin temperature at the beginning of each test session. The thermode area of 255 mm^2^ balances the psychophysical requirement for reliable detection of temperature change against the system requirement of delivering a power-efficient hardware package which does not require active cooling.

The advantages of packaging haptic and thermal stimulation together are two-fold. Firstly, it makes the SenseCheQ test protocol more streamlined, as the participant need not reposition their hand part-way through testing. Secondly, and more importantly, simultaneous thermal and haptic control allows the thenar eminence to be held at a stable temperature of 32 °C during vibration detection testing. This is important because vibration detection is known to be influenced by skin temperature, with a marked decrease in sensitivity as skin temperature falls^31–34^. Skin temperature varies greatly depending on environmental factors, activity levels, circadian rhythms and autonomic dysregulation, all of which can be affected in people with cancer, undergoing chemotherapy^35,36^. This integration of thermo-haptic stimulation is unique; there is no equivalent commercially available device or experimental equipment that affords this capability.

Vibration and thermal ramping profiles were defined to balance test duration against detection-threshold reliability. Our final design parameters are summarised in Table 1.

**Table 1.** Design parameters for SenseCheQ.

| Design Parameter | Parameter Requirement |
| --- | --- |
| Test site | Hand (thenar eminence) |
| Thermode Area | 255 mm<sup>2</sup> |
| Thermal Ramp Rate | 1°C/s |
| Haptic (Vibration) Frequency | 128 Hz |
| Haptic Amplitude Range | 0 – 15 µm |
| Patient response interface | Haptic mechanical push buttons |
| Instruction display | Adequate to allow large text |
| Maximum session duration | 15 mins |

### Calibration Routine

To further reduce variance in vibration detection threshold measurements, SenseCheQ employs a haptic-calibration routine (Fig 3A). This was developed to more reliably deliver a predefined target vibration amplitude envelope in each test session, reducing the influence of sources of variance. The amplitude of vibration stimulus delivered to the user’s thenar eminence depends on both the amplitude of the drive signal passed to the haptic actuator and the force applied to the SensePad by the participant’s hand allied to the coupling friction. Compressive (downward) force on the stack leads to mechanical damping, resulting in reduced mechanical vibration amplitudes at the SensePad. To account for this variability, an uninterrupted full-range vibration ramp is initially delivered to the participant. During this calibration ramp, the accelerometer measures the movement of the SensePad and the amplitude of vibration is computed. Subsequently, a regression analysis is used to determine the within-test session relationship between haptic actuator command drive values and SensePad amplitude of vibration at each stage of the calibration ramp. The regression model is then used to calculate correction factors for the haptic drive values at each step in the subsequent test ramps.

**Fig 3.**
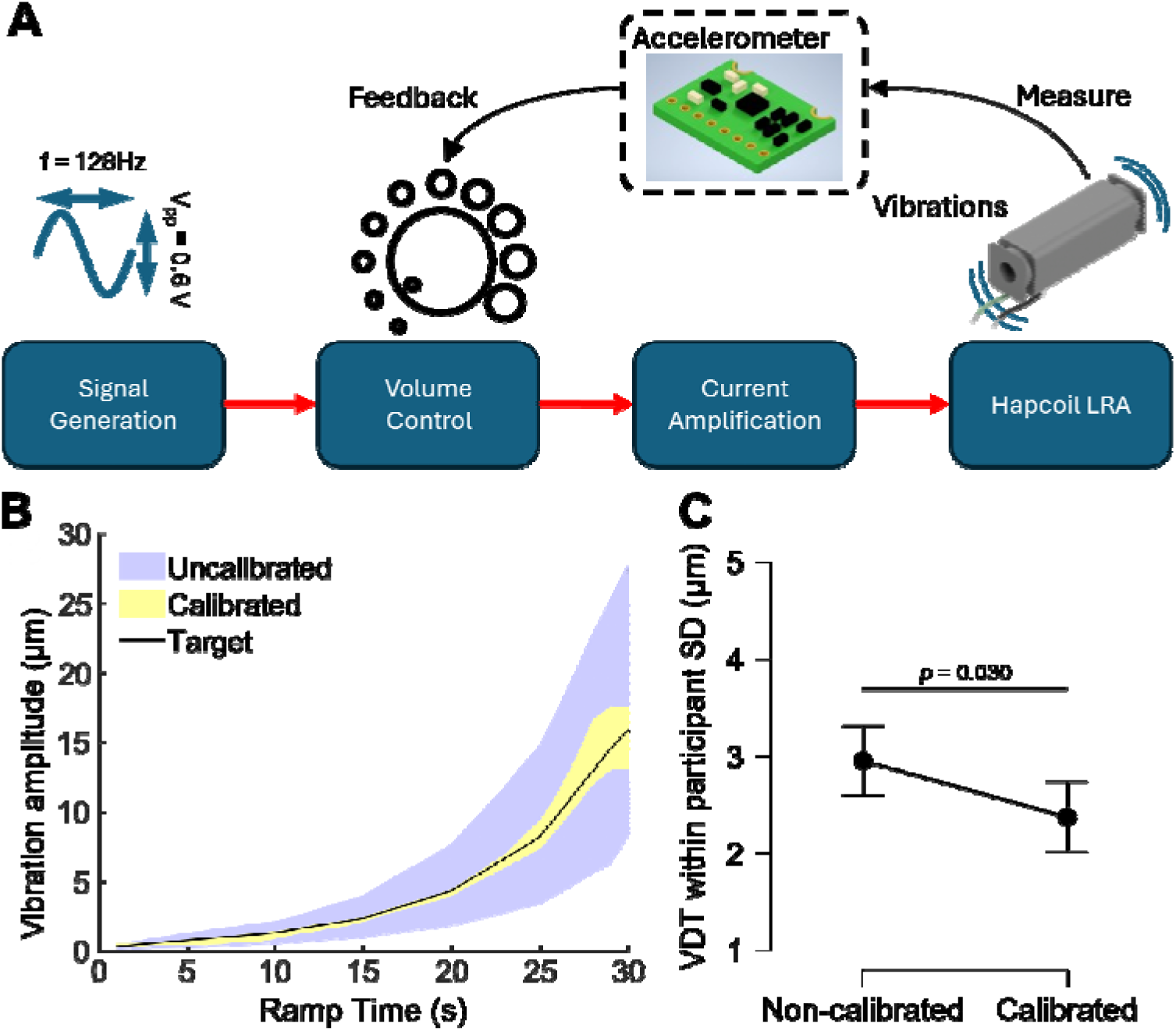
SenseCheQ Vibration Stimulation. A – block diagram schematic of the haptic drive circuitry. The accelerometer provides feedback for the vibration stimulation to allow calibration and subsequent monitoring of the haptic ramps delivered to SenseCheQ users. B – demonstration of the effect of haptic calibration on the envelope of delivered vibration ramps at 128 Hz stimulation frequency. Ten ramps while applying different amounts of pressure were delivered with and without calibration. The calibration routine reduces the amplitude envelope of vibration stimulation to lie closer to the desired set-point values. Shadings represent min-max ranges for calibrated and uncalibrated ramps C – plot demonstrating the effect of the haptic calibration routine on vibration detection thresholds at the volar forearm of healthy participants. Within participant variance is significantly reduced when a haptic calibration routine is used (paired t-test; *t*(9) = 2.58, *p* = 0.030, *d* = 0.82 [0.08, 1.52]).

By comparing calibrated and non-calibrated vibration ramps, we find that the calibration routine reduces the variance of the haptic stimulus delivered to the user by 80% [74.89, 85.96] (Fig 3B) when coefficients of variation are compared across ramp steps (Z = 4.78, *p* < 0.001). Compared to the target ramp, the average deviation from target amplitudes is reduced by 81.23% [76.01, 86.45] (see Methods for details). The increased precision of the physical stimulus being delivered reduces the within-participant variance across repeated measurements of vibration detection threshold (Fig 3C). This increased reliability improves the ability of SenseCheQ to detect subtle changes in perception associated with early CIPN.

### Thermal Performance

SenseCheQ was designed to deliver heating and cooling ramps at a rate of 1°C/s from a skin temperature clamped at a baseline of 32°C. Fig 4 displays typical cooling and heating ramps. The Proportional-Integral (PI) controller constants for SenseCheQ are set to operate under the assumption that a user’s hand is in contact with the SensePad throughout the duration of the test, acting as a thermal mass. Performance testing shows reliable compliance to the targeted ramp rate (Fig S1A) and stable maintenance of the baseline temperature at 32°C (Fig S1B & S1C) between thermal ramps across 192 sessions conducted at home by 16 users under different conditions and on multiple devices.

**Fig 4.**
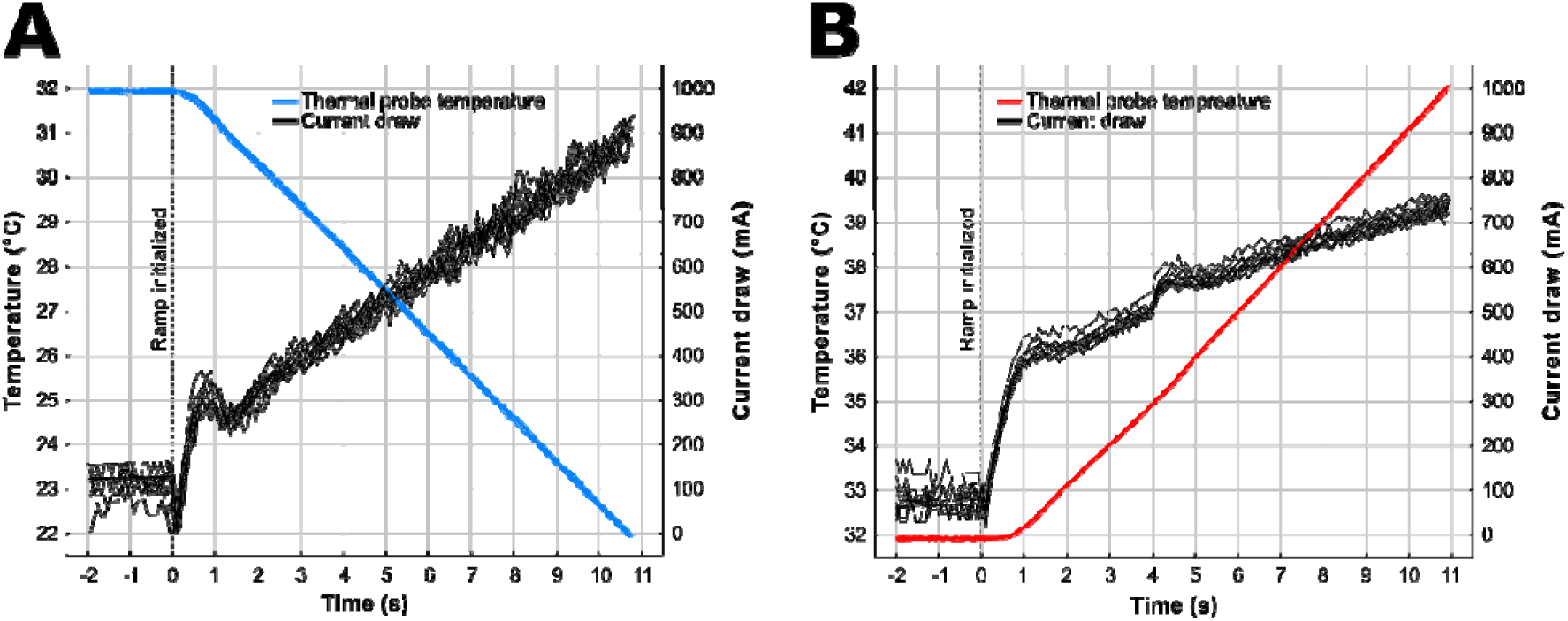
Typical thermal ramps delivered by SenseCheQ. The figure shows 10 consecutive thermal ramps (on a single user) overlaid for cooling (A) and heating (B) with the first 2 seconds at baseline (32 °C) preceding the ramps. Both cooling and heating follow stable 1°C/s gradients. Current draw through the Peltier during the ramps is plotted against the right-hand axes and reveals little variance, most notably no escalation of current draw due to repeated cooling or warming which reflects ample heat-sinking capacity and thermal dissipation within the device.

### Home-based testing in Healthy Participants

To assess the performance of SenseCheQ outside of a controlled laboratory setting we conducted an experiment in the home setting. Healthy volunteers (N=30) completed an initial familiarization session in an environmentally controlled clinic room followed by three self-administered home test sessions. In each session, users completed the standard protocol (from Fig 1A) with the average session lasting 7 minutes. The first two home sessions were used to assess test-retest reliability, whilst the final session employed a modified protocol in which skin temperature was not clamped prior to and during VDT measurement, to determine whether there was an advantage of thermal control of baseline skin temperature.

### Test-Retest Reliability

To establish test-retest reliability, we compared Home Session 1 and Home Session 2, conducted approximately 24 hours apart, on all three test parameters. VDTs were not significantly different between the two sessions (*M*_difference_ = 0.02 µm [−0.02, 0.04]; *t*(29) = 0.59, *p* = 0.594, *d* = 0.10) (Fig 4A). The mean absolute difference between the within subject measurements was 0.16 µm [0.09, 0.23]. Considering that differences between the two sessions were small and that the range of thresholds is narrow as testing was conducted on healthy participants at a sensitive area (thenar eminence), we evaluated test-retest reliability by computing Spearman rank-correlation coefficients (ρ(28) = 0.75 [0.53, 0.87], *p* < 0.001) and intraclass correlation coefficient (ICC(2,1) = 0.66 [0.41, 0.82]) indicating that VDT measurements were reliable in the home setting (Fig 5A).

**Fig 5.**
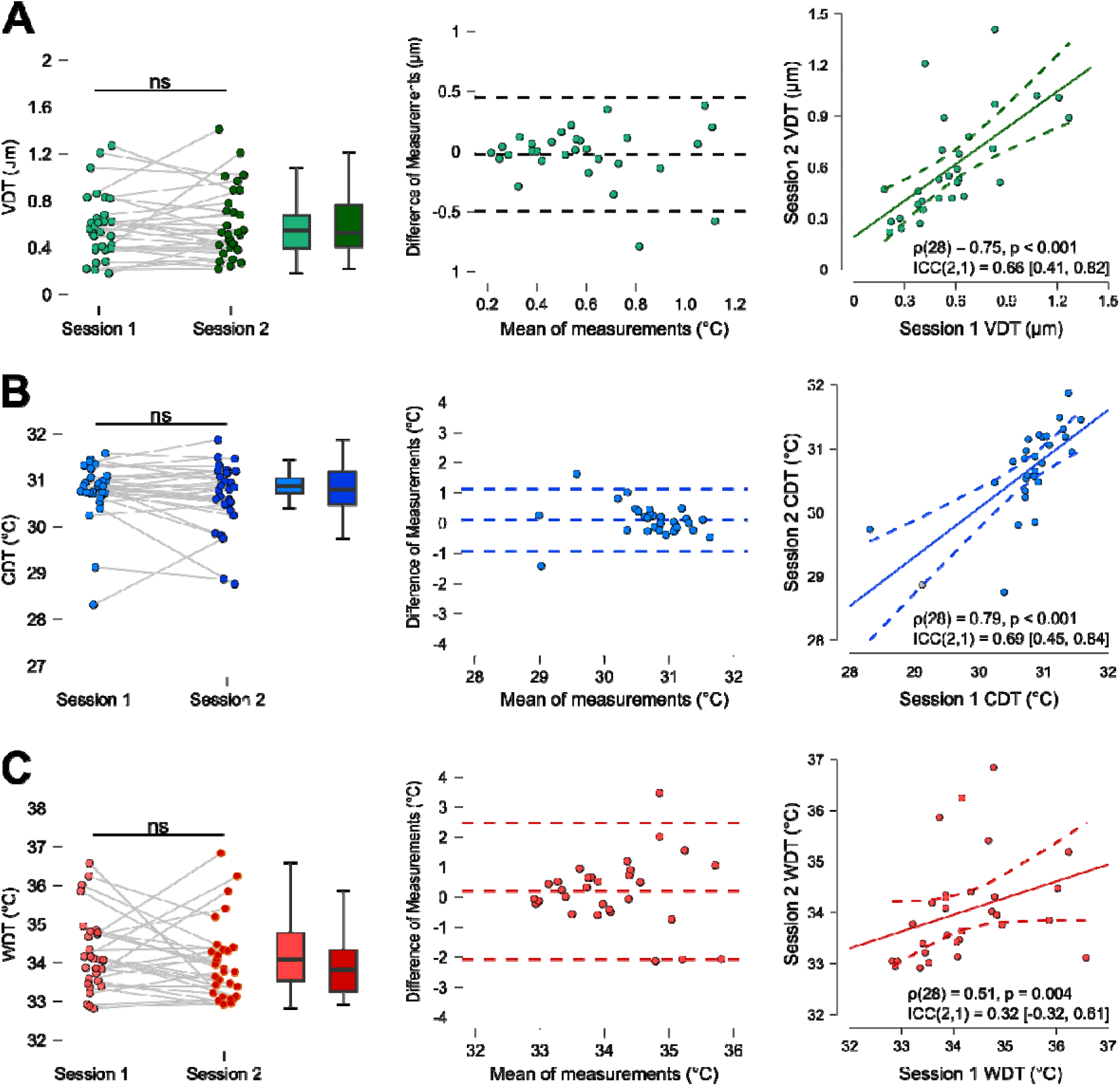
Test-retest reliability analysis. A – Vibration detection thresholds. B – Cold detection thresholds. C – Warm detection thresholds. Columns show individual data and boxplots from the two sessions (left); Bland-Altman plots of difference between sessions in which dotted lines represent mean difference and 95% CI (middle); scatterplots with linear model fit, Spearman’s coefficient and ICC between the sessions (right). There were no group-level significant differences between the sessions.

Similarly, for thermal testing, the CDTs were not significantly different between the two sessions (*M*_difference_ = 0.10°C [−0.09, 0.30]; *t*(29) = 1.07, *p* = 0.293, *d* = 0.20) and the mean absolute difference was 0.38°C [0.23, 0.52]. Test-retest reliability was good as measured with Spearman’s rank correlation ρ(28) = 0.79 [0.60, 0.90], *p* < 0.001 and ICC(2,1) = 0.69 [0.45, 0.84]. WDTs were also not significantly different between the two sessions (*M*_difference_ = 0.20°C [−0.24, 0,63]; *t*(29) = 0.93, *p* = 0.363, *d* = 0.17) and the mean absolute difference was *M* = 0.85°C [0.55, 1.15]. Test-retest reliability for WDT was relatively weaker with a Spearman’s rank correlation of ρ(28) = 0.51 [0.18, 0.74], *p* = 0.004 and ICC(2,1) = 0.32 [−0.04, 0.61].

### Advantage of Skin Temperature Clamping

To determine whether controlling skin temperature during VDT measurement is beneficial in reducing variance in measurements we compared the within subject measurements made with and without skin temperature clamping (home session 2 vs home session 3, respectively) showing a larger co-efficient of variation in the unclamped session (Fig 6A). We also assessed the relationship between the differences in VDT between the unclamped and clamped sessions with the skin temperature deviation in the unclamped session from the normal clamped baseline (32°C). This was based on the expectation that participants with cooler skin in the unclamped session would have lower sensitivity to vibration. This was indeed the case (Fig 6B); participants with cooler skin temperature had larger VDT deviations in Home Session 3 when compared to Home Session 2 (*r*(28) = −0.37 [−0.64, −0.01], *p* = 0.046).

**Fig 6.**
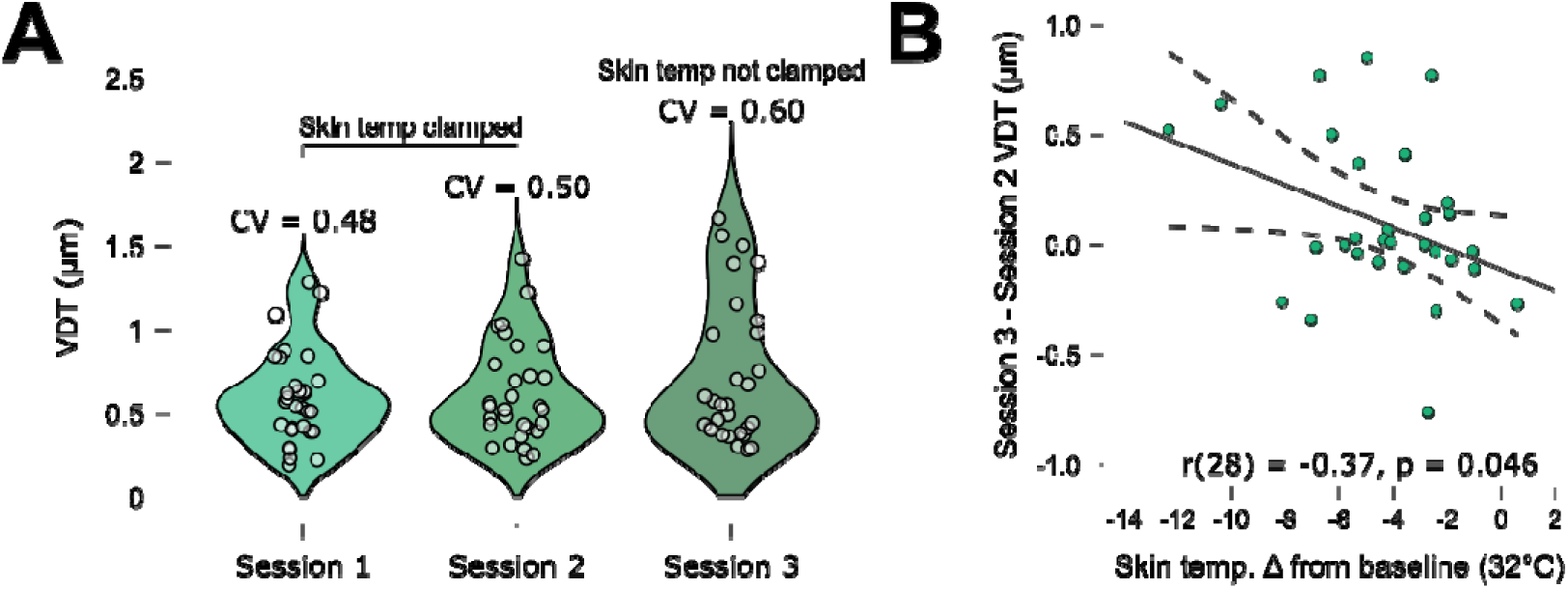
Impact of clamping skin temperature on vibration detection thresholds. A – violin plots of the three home sessions demonstrate increased variance of VDTs in Home session 3 in which skin temperature was not clamped during VDT measurement (CV = coefficient of variation). B – VDT change as a function of skin temperature deviation from the usual clamped baseline (32°C). Skin temperature deviations predict VDT differences when compared to Session 2 in which skin temperature is clamped to the baseline of 32°C during measurement.

### Patient Home Testing Case Studies

Following the healthy participant home study, we collected pilot data using SenseCheQ from a small group of patients. We show data collected from two female breast cancer patients, recruited prior to starting chemotherapy treatments and self-tested regularly during their chemotherapy treatment. The first patient received a taxane-based neurotoxic agent, and the second patient received two neurotoxic agents in combination; one taxane and one platin-based. A supervised SenseCheQ test session was completed in the lab, prior to and post-chemotherapy. In between, patients conducted home testing sessions with SenseCheQ on a weekly basis. It is worth noting that very good compliance with the testing schedule has been observed with the large majority of patients completing the home testing study to date (N=13/20). Patients have completed 83 of the 99 planned weekly home testing sessions (84%). Patient 1 reported no CIPN-related symptoms measured using questionnaires (Brief Pain Inventory, EORTC CIPN20, DN4) and assessment (TNSc) during visits or retrospectively throughout their treatment (6/12 weekly cycles completed as of their second lab visit). Longitudinal testing with SenseCheQ showed no changes in sensory function across this period (Fig 7A). Patient 2 was on a combined regime with infusions every three weeks, and they conducted home SenseCheQ sessions every week. This patient reported paraesthesia (tingling) in their fingers in the period following each infusion which recovered just before the next infusion cycle. They also reported fewer symptoms following the fourth infusion (before their second lab test session). We recorded a 14-point increase in their CIPN20 scores and a 3-point increase in their TNSc assessment between start and end of their treatment period indicating the development of acute neuropathy. The SenseCheQ testing mirrors this cyclical pattern of paraesthesia symptoms as VDT becomes significantly worse two weeks after each infusion cycle for the first three cycles but not the final infusion prior to their second lab visit (Fig 7B). These acute sensory changes did not trigger a change in their treatment.

**Fig 7.**
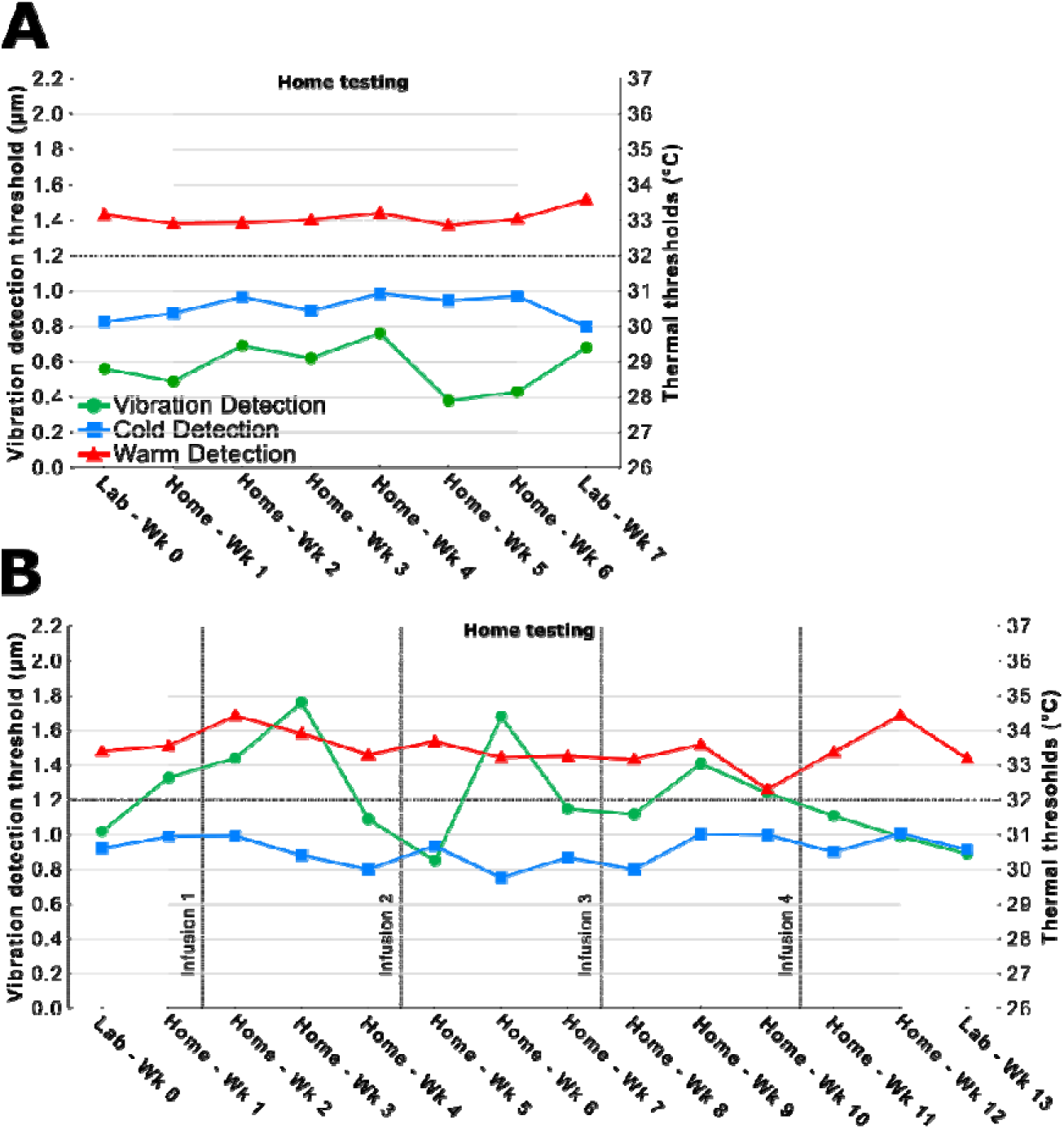
Longitudinal testing with SenseCheQ by patients during chemotherapy. A – Patient 1 detection threshold results assessed up to half-way through their treatment (6/12 weekly infusions) with no change to sensory function. B – Patient 2 testing up to infusion 4/6 of their chemotherapy treatment. Cyclical changes in VDT correspond to symptoms of paraesthesia in the fingers following chemotherapy infusions.

## Discussion

The aim of our project was to develop reliable, user-friendly sensory testing, which can be self-administered by patients in their homes (i.e. autonomously) during their chemotherapy treatment. Enabling regular sensory testing is crucial, as it allows rapid detection of small, but potentially clinically meaningful trends in sensory function that may aid timely clinical decision making before irreversible nerve damage occurs. This could lead to better informed decisions on treatment modification assayed quantitatively against repeated sensory function testing. Conversely this approach, by providing quantitative evidence of functional recovery after each cycle of treatment, could also reduce the risk of more radical treatment modifications such as complete cessation, which would then be less likely to adversely impact cancer outcomes.

We have demonstrated the ability of SenseCheQ to deliver reliable sensory stimuli in healthy volunteers and patients undergoing chemotherapy for cancer. This has been achieved through technological innovations to reduce variance from environmental and user-related sources. Calibration of the vibration stimuli and control of skin temperature during VDT measurement increased the reliability of both the physical stimuli being delivered and the participants’ detection thresholds. These key innovations improve the confidence to detect smaller changes in stimuli (at a sub-micron precision) over time during treatment. Beyond objectively measured performance and reliability, we also consistently received encouragingly positive feedback from both healthy participants and patients.

In addition, cost-effectiveness has been an important consideration throughout the development of SenseCheQ to facilitate widespread adoption and home deployment. We have accordingly used off the shelf componentry wherever possible and limited its specification to a core test feature set. At this point, the component cost per device has been kept to <£500 (<$670) which compares relatively favourably with the equivalent commercially available QST equipment which is typically more than an order of magnitude more expensive.

We have demonstrated good test-retest reliability using three inter-related but independent indicators. The mean differences between two home test sessions are non-significant, with no evidence of systematic bias over time. The lack of any significant training effects speaks to the ease of use and quick familiarization with the protocol. Test-retest reliability, as measured by rank order and intraclass correlation coefficients between the two home sessions, revealed good reliability for VDT and CDT but lower reliability for WDT. Thermal detection thresholds assessed using formal QST testing in a controlled laboratory environment are usually reported to have fair to good but not exceptional reliability^37^. For example, Moloney et al.^38^ reported fair to good CDT and WDT reliability (ICC ∼0.4) when measuring thresholds at the hand in a healthy cohort. SenseCheQ achieves considerably better reliability for CDT (and VDT) with slightly lower ICC for WDT.

Reassuringly, the absolute magnitude of the differences between two home sessions are small. These session-to-session differences are much smaller in magnitude than the expected pre-post chemotherapy differences in patients. In studies informing the development of SenseCheQ^20^, we found much larger differences between patients with confirmed CIPN and healthy control participants than the session-to-session differences noted using SenseCheQ at home. For CDT, the average difference between healthy controls and patients with CIPN was 4.1°C^20^ whereas the largest session-to-session difference observed in the home study was 0.8°C. Similarly, the difference in WDTs between healthy controls and CIPN patients was 2.6°C on average^20^, which is larger than the session-to-session difference observed in this SenseCheQ home study. A meta-analysis of pre-post chemotherapy differences in VDTs revealed an average change from 0.8 µm at baseline to 2.6 µm post-chemotherapy^39^. This is a considerably larger change than any session-to-session difference recorded in the SenseCheQ home study which is able to resolve sub-micron changes in vibration detection. On this basis we anticipate being able to track trajectories of change in sensory function over time that reflect neurotoxicity early in its pathogenesis.

We have been able to collect for the first time patient data from regular, self-administered home sensory testing during chemotherapy. This early data indicates good compliance with SenseCheQ testing through regular use by people going through physically and mentally demanding chemotherapy treatments. This data also shows evidence of alignment between nerve function measurement with SenseCheQ and clinical symptom report during treatment. It is particularly relevant to note that there is evidence that SenseCheQ measurements track sensory changes (paraesthesia) that are considered an early sign of the development of acute neuropathy but at a point when it has not triggered changes in treatment regimens, has not caused serious impairment and was not expected to develop into persistent issues. This increases our confidence going forward that SenseCheQ will be suitable for tracking clinically meaningful changes and inform decision making. It is also clear that the sensory profile of the second patient case study (Fig 7B) demonstrates that testing more often is preferable. If the patient had used the device just on days prior to infusions, the cyclical changes in VDT with each dose of chemotherapy would not have been detected, potentially missing evidence of early nerve toxicity.

The results from these studies provide clear directions for further work including iterative design evaluation for optimised ergonomics (e.g., ambidextrous design), increased accessibility options (e.g., multiple languages) and on-device collection of patient reported outcomes synchronous with the sensory testing. We aim to assess the benefit of incorporating a Peltier with a larger total surface area and decrease ramp rates, both of which may increase sensitivity and reliability of WDT measurements^41–44^. However, this has to be counterbalanced by the potential advantage of smaller thermal probe size for detecting loss of nerve function due to CIPN^20^. Similarly, there needs to be caution over reduction of ramp rates as accommodation effects may reduce validity^44^ at very low rates. We also plan to develop our haptic calibration approach to take account of small dynamic fluctuations in the applied hand pressure during a test session. To provide further context for the adoption of home sensory testing in standard clinical care, there is a need for qualitative studies with patients and clinicians as well as healthcare economics modelling to identify obstacles and opportunities. While we have demonstrated the feasibility and reliability of self-administered home-based QST but longitudinal work at scale will be required to properly evaluate the predictive utility of QST for early detection of chronic CIPN.

In conclusion, we have developed a cost-effective, user-friendly, reliable, self-administered sensory-testing device and protocol focused on detection of CIPN. We note that SenseCheQ could also be a valuable tool for community-based longitudinal monitoring of a range of other neuropathies including Diabetes-induced peripheral neuropathy^45^. Similarly, it may provide a surrogate biomarker of disease status against which novel therapeutics could be assayed.

## Methods

### SenseCheQ hardware specification and operation

#### SenseCheQ Hardware

The components required to deliver and monitor the stimuli are housed below the SensePad and can be seen in the exploded view of the “Stack” (Fig 1D). Working from bottom to top this comprises:

- Linear resonant actuator (LRA) haptic device (Actronika, Hapcoil One), which provides vibration stimulation.
- Mounting bracket, a bespoke 3D printed component which houses the LRA.
- Heatsink (Malico, MBH42.5002-25P/2.6).
- Peltier device (12.3 mm x 12.3 mm x 3.4 mm, 4.5 W, 1.54 Ω) (Tark, 71035-506) which provides thermal control and stimulation.
- “stack top” which is a custom-designed 3D printed component.
- bespoke flexible printed circuit board – which provides electrical connections to the Pt1000 thermometers (1^st^ Innovative Sensor Technology, P1K0.232.6W.A.007)
- 15-bit, 800Hz accelerometer (Adafruit/NXP Semiconductors, MMA8451Q) which is used to monitor the vibration stimulation delivered to the participant in real-time informing the SenseCheQ haptic stimulation calibration routine, housed within the stack top – which is EMF-shielded to ensure the integrity of small-level analogue signals.
- aluminium face-plate (15 mm x 15 mm) which acts as the thermal contact point of the SensePad; the three Pt1000 thermometers which are used for measuring skin temperature, informing the thermal (PID) control, and as the sensor in the independent thermal cut-out safety hardware.

The control system comprises a Raspberry Pi 4 (4 Gb) and two Arduino microcontrollers. The Raspberry Pi acts as the master-controller, taking inputs from the response buttons, displaying test-prompts via the TFT screen, and directing the behaviour of the microcontrollers. The thermal and haptic control are managed separately by two Arduino microcontrollers (MKR Zero and Nano ESP32 respectively). The Raspberry Pi is used to coordinate operation and run the SenseCheQ graphical user interface (GUI), whereas microcontrollers are used for real-time control of thermal and haptic stimulation modes thereby minimising issues with timing critical processes. To minimise the risk of thermal run-away the thermal control was given a dedicated microcontroller with a layer of software temperature cut outs (secondary to hardware cut outs).

Power is provided to the SenseCheQ device via a 60601-1 certified AC/DC converter, to ensure that there is electrical isolation between the participant and mains power. This plugs in to the rear left-hand corner of the SenseCheQ enclosure via a 1.6 mm barrel-jack socket.

#### SenseCheQ Software

Standard Arduino sketches (requiring no custom or purpose-made libraries) contain the code which controls microcontroller behaviour during calibration, vibration and thermal ramps and during measurement of initial skin temperature, ambient temperature and amount of pressure being put onto the SensePad. A Python script deploying standard libraries (such as numpy, scipy, sklearn, pandas, signal and gpiozero) serves as the central controller. It is used to initiate microcontroller behaviour, record and store data, execute calibration-related data transformations and computations as well as deliver the user interface (built using the guizero library). The Python script is executed by the Raspberry Pi running a Bookworm distribution of the Pi OS (2024-10-22-raspios-bookworm-arm64).

#### Peltier Drive Circuitry

Peltier devices are current-controlled ^46–48^, i.e. their heating or cooling power is controlled by the current drawn through the Peltier device. The voltage-current relationship for a Peltier device is not constant – it changes depending on the thermal gradient across the Peltier device, and therefore a current source is used for Peltier device control^49^. In SenseCheQ, the Peltier is driven by a programmable current sink, which was designed based on the work of Bradley et al.^50^ (Fig S2A). A zero-drift precision operational amplifier (op-amp) (Texas Instruments, OPA189) receives a variable DC voltage from the thermal microcontroller (Arduino, MKR Zero) to its non-inverting input (V_Iset_). The inverting input receives a feedback voltage from the measurement node at the top of a reference power resistor. The output of the op-amp is connected to the base of a high-gain Darlington Pair (STMicroelectronics, BDX33C), which modulates the current flowing through the Darlington Pair, and therefore through the entire current path, such that the voltage at the control node is equal to V_Iset_. A 1 Ω reference resistor is used in this case, to provide a one-to-one ratio between V_Iset_ and the current draw through the Peltier device, as well as to ensure that sufficient voltage-overhead remains to avoid voltage-clipping across the Peltier device. The Peltier device is connected between the current sink and the 12V / 5A supply via H-Bridge circuitry (2 x AP331A comparators, 2 x DMN3023L N-FETs, 2 x DMP3096LQ P-FETs), which provides control of the current direction through the Peltier device. Current draw is determined by a proportional-integral (PI) control algorithm, which runs continuously on the thermal control microcontroller. Feedback for the PI control algorithm is provided by one of the Pt1000s attached to the thermal face plate, which is polled by the microcontroller every 100 ms via a Pt1000 signal amplifier (Analog Devices, MAX31865).

#### Thermal Safety

Because SenseCheQ is designed to be used by patients at home with no supervision, safety is a vital concern. The biggest risk is thermal malfunction, which in a worst case could lead to skin burns, especially in a patient with sensory loss. To mitigate against this risk, there are triple safeguards in place to prevent thermal run-away. The first line of defence is a software cut-out, which sets the current draw through the Peltier to zero if 43°C is exceeded by the device during operation. In addition to the software cut-out, a hardware cut-out (Fig S2B) will force-shutdown of the thermal circuitry if the faceplate exceeds 45°C. In this circuitry, a comparator (Diodes Incorporated, AP331A) monitors two legs of a Wheatstone Bridge, with the reference leg set to 45°C equivalent resistance via the adjustment of a trim-pot. The other leg comprises a dedicated Pt1000 mounted on the thermal face plate plus a set resistor to create a potential divider. If the safety Pt1000 measures a temperature greater than 45°C (in other words, the potential read at the node above the Pt1000 exceeds the potential on the corresponding node on the other Wheatstone Bridge leg), the comparator will drop its output low, which forces the thermal control Arduino into reset status, thereby shutting down the thermal control circuitry by forcing the current sink control voltage to 0 V. Finally, in the consequently unlikely event that complete control is lost, resulting in a short circuit between the Peltier and ground, a 2 A slow-blow fuse is placed in series with the Peltier which will blow quickly under the maximum current draw of 5 A. It is worth noting that no thermal cut-out events have been observed under standard test conditions, which enumerates many thousands of thermal ramps, spread across hundreds of SenseCheQ test sessions, with different multiple SenseCheQ devices, and in many different environmental conditions.

#### Haptic Drive and Feedback Control

The haptic stimulation is managed by a dedicated microcontroller (Arduino Nano, ESP32). A block-diagram schematic of the Haptic drive circuitry is displayed in Fig 2A. Rather than generating a sine wave directly from the Arduino, a signal generation module (Analog Devices, AD9833), paired with a 1 MHz crystal (Abracon LLC, ASDMB-1MHz), is used to generate a 0.6 V_pp_ sine wave at 128 Hz to match the Rydell Seiffer tuning-fork frequency typically used in clinical VDT assessment. An active amplifier/filter sub-circuit is used to boost the signal to 1.8 V_pp_ and filter out the high-frequency elements generated by the signal generation module during wave-writing. The signal is passed through a 12-bit digital potentiometer (Analog Electronics, AD5142A), which acts as volume control, before the signal is current-amplified by a 4 W class-AB audio amplifier (Texas Instruments, LM4990). This current-amplified signal drives the haptic LRA located at the bottom of the stack producing vibration in a lateral plane. The 3-axis accelerometer, housed in the top of the stack, provides real-time measurement of the vibrational stimulation that the participant is receiving as well as providing the feedback signal for the Haptic calibration routine.

#### SenseCheQ Haptic Calibration Routine

Once the user is positioned for their test session, with their hand resting comfortably on the SensePad, a full-range, uninterrupted, stepped haptic ramp is delivered via the stack. The vibration ramp is consists of 30 steps, each lasting for a second. The choice to generate a stepped ramp was made so that enough accelerometer data could be gathered at each step to reliably compute displacement. Additionally, during measurement, this stepwise stimulation minimizes the impact of response times on measured thresholds, as participants have a second to detect and respond at each step. After the calibration ramp, a regression model is fitted to determine the relationship between drive values of the vibration circuit and displacement (amplitude) delivered across the ramp (30 data points). The resulting regression model is then used to predict what the drive values should be to deliver pre-determined target drive values across the ramp. The predicted drive values are then used for the subsequent VDT measurements in the test session. Calibration is conducted once per session, but the accelerometer data is collected throughout every ramp and stored on device for metrology and later analysis.

Evidence of more consistent delivery of vibration stimulation following calibration can be found in Fig 3B. The data was acquired by applying variable pressure to the SensePad and running an uncalibrated and calibrated vibration ramp at each pressure level. On half of the pressure levels the uncalibrated ramp preceded the calibrated one and in the other half it was the opposite. Coefficients of variation were computed across pressure levels at each drive value across the 30-second vibration ramp both for uncalibrated and calibrated ramps. The coefficient variations of uncalibrated and calibrated ramps were then compared using a Wilcoxon signed-rank test which revealed less variation in calibrated ramps. Additionally, absolute differences in displacement between each ramp and the target stimulus envelope were computed at each step of each ramp. These deviations from target were then compared between uncalibrated and calibrated ramps revealing much better adherence to target after calibration.

#### Enclosure, screen and user interactions

The stack is enclosed in a 3D printed PLA case of dimensions 250 mm x 200 mm x 60 mm. The screen is situated on the top face of the box, offset to the right mounted in an angled bezel (5” thin film transistor (TFT) screen (Waveshare, WAV-18396)). This location was selected so that a majority of people would be able to place their non-dominant hand on the SensePad and use the interaction buttons with their dominant hand (although there is no need in principle to limit testing to the non-dominant hand). Below the touch screen are the two tactile interaction buttons (RS, 820-7561 (Green), 820-7568 (Red)) which participants use to progress through the sensory test protocol and indicate their detection of the test stimuli. In the top left of the case there are two LEDs. One indicates that power is supplied to the device (red) and the other indicates that the Raspberry Pi is on (blue).

### Assessment of the influence of calibration for VDT measurements

#### Participants

Ethical approval was obtained from the University of Bristol Research Ethics Committee (project ID:9994). Participants (N = 10; 4 female; M_age_ = 30.90, Min-Max = 21-58) were recruited among University of Bristol students and staff. Exclusion criteria were acute or chronic pain conditions; regular or recent (within 24 hours) analgesic use; presence of neuropathy or other neurological impairment that may affect sensory function; broken or infected skin at the testing area, or allergies to latex and alcohol. Participation was completely voluntary without reimbursement.

#### Materials

We measured VDT using only the vibration testing circuit of the SenseCheQ test device. For this purpose, we used a peripheral enclosure containing the haptic actuator which could be affixed to the volar forearm. The forearm was chosen for this experiment due to its lower sensitivity and larger between-participant variance than the thenar eminence. It was, therefore, more likely to reveal an effect of the calibration on detection thresholds when testing is done with healthy participants.

#### Procedure

The haptic enclosure was affixed to the volar forearm, and vibration detection threshold measurements were conducted with and without the calibration routine prior to testing. The peripheral was then removed, affixed again after a short break, and the measurements repeated. Half of the participants started with calibrated and half with non-calibrated ramps in a counterbalanced design (participants were unaware of allocation). There was a total of 8 ramps per condition in each measurement.

#### Analysis approach

To determine whether calibration increased reliability, we computed standard deviations across all non-calibrated and all calibrated trials for each participant. Following assumption testing a repeated measures t-test was conducted to compare variance in the non-calibrated and calibrated conditions.

### Healthy participant at home study

#### Participants

Ethical approval was obtained from the University of Bristol Research Ethics Committee (project ID:9994). A total of 30 participants (24 female; M_age_ = 37.10, SD = 11.80, Min-Max = 23-64) were recruited among University of Bristol staff. Exclusion criteria were the same as for the *VDT* measurement studies. Participants were reimbursed £20 upon completing the study.

#### Materials

All testing was conducted using the SenseCheQ device (as described in the Results and Methods sections). The standard testing protocol can be seen in Fig 1D. Each sensory testing session started with measurements of skin and ambient temperature. The stimulated area (thenar eminence of the left hand) was then clamped to 32°C prior to a vibration calibration routine. This brief setup phase was followed by the testing protocol itself in which VDTs were measured first, followed by CDTs and WDTs. Each of the thresholds were measured four times per session; following the initial calibration ramp, VDT was measured as the amplitude at which participants first detected the SensePad (Fig 1B & C) start vibrating (at a frequency of 128 Hz). CDT and WDT were measured as the temperature at which participants first noticed cooling and warming from the baseline temperature of 32°C. Each session took about 7 minutes to complete; participants also completed a feedback questionnaire (available on **<u>OSF</u>** alongside data).

#### Procedure

Participants attended the familiarization session at the NIHR Bristol Clinical Research Facility with ambient temperature of 21°C. After confirming eligibility and providing written consent, participants were briefed and completed an initial test session and received the device for home testing. The remaining three sessions were conducted by the participants at their homes with the only requirement being that sessions had to take place on three separate days. Participants were told to conduct the sessions when they had time to focus undisturbed. On average, the four (one in lab and three at home) sessions were completed within seven days with a minimum period of 18 hours between any two sessions.

In a normal test session, skin temperature of the thenar eminence is clamped to a baseline of 32°C before any of the measurements commence. In the final home session, skin temperature stabilization occurred *after* VDT measurement. This was done to determine whether there is an advantage (more stable measurements) of controlling skin temperature when measuring VDT.

#### Analysis approach

VDT, CDT and WDT for each participant in each session were computed as medians across the four trials. After assumption checks, repeated-measures t-tests and bivariate linear regressions were conducted to determine differences between sessions and test-retest reliability as well as the relationship between skin temperature and VDT.

### Patient case studies

#### Participants

Ethical approval was obtained from Health Research Authority and Health and Care Research Wales (IRAS project ID: 327541). Two breast cancer patients (50-60 years old) were recruited from the Bristol Haematology and Oncology Centre, University Hospitals Bristol and Weston. Patients were referred to the study by their clinical care teams after being scheduled for potentially neurotoxic chemotherapy regime. After initial contact patients were given further information (via participant information sheet) and eligibility was established (same as in the healthy participant study). Patient 1 received a taxane-based chemotherapy regimen (12 cycles weekly). Patient 2 received the 6 cycles once every 3 weeks of a combined taxane and platin-based regimen.

#### Materials

The TNSc^51^, EORTC QLQ-CIPN20^40^, Brief Pain Inventory^52^ and the Douleur Neuropathique 4 questionnaire^53^ were administered during the lab sessions. Lab visits took place prior to patients starting their chemotherapy treatments and upon returning the SenseCheQ device which took place past the mid-point of their scheduled treatments.

#### Procedure

After arriving at the NIHR Bristol Clinical Research Facility, eligibility was confirmed and written consent obtained. atients were familiarized with the SenseCheQ testing equipment and conducted a test session with the device before completing the questionnaires and taking the device with them for home testing. Both patients conducted self-administered home testing on a weekly basis, the day before their chemotherapy infusion on weeks in which they were to receive chemotherapy; at the point they felt at their best physically and psychologically with the most rest between infusions. This also allows for any acute, non-neuropathic, impact on measurements to minimize following an infusion. Patient 1 attended their second visit halfway through their treatment and Patient 2 did so after completing 2/3 of their scheduled infusions, during which the same procedure from the first visit to the research facility was repeated. Patients were reimbursed at a rate of £10/h for the lab visits and £5 per completed home testing session.

## Data Availability

All data produced in this manuscript is available via Open Science Framework (https://doi.org/10.17605/OSF.IO/4GD2H).

## Acknowledgments

Special thanks to our patient partner group who have contributed to each step of SenseCheQ development and testing programme, (Alan Young, Iain MacLeod, Rubina Zafar, and the other members of the group who have elected to remain anonymous).

The research was carried out at the National Institute for Health and Care Research (NIHR) Bristol Clinical Research Facility (CRF). The views expressed are those of the author(s) and not necessarily those of the MRC, Arthritis UK, Eli Lilly, the NIHR or the Department of Health and Social Care.

The SenseCheQ research project was funded as part of the Advanced Pain Discovery Platform by a consortium made up of UKRI MRC, Arthritis UK, and Eli Lilly all coordinated by the MRC [MR/W027925/1] as well as by UKRI EPSRC [UKRI1437]. BT was also supported by an APDP grant as part of the Partnership for Assessment and Investigation of Neuropathic Pain: Studies Tracking Outcomes, Risks and Mechanisms (PAINSTORM) consortium [MR/W002388/1].

Data is available via OSF repository (https://doi.org/10.17605/OSF.IO/4GD2H).

