## Supplementary materials for "SenseCheQ: Home-based Nerve Function Self-Assessment using Autonomous Quantitative Sensory Testing"

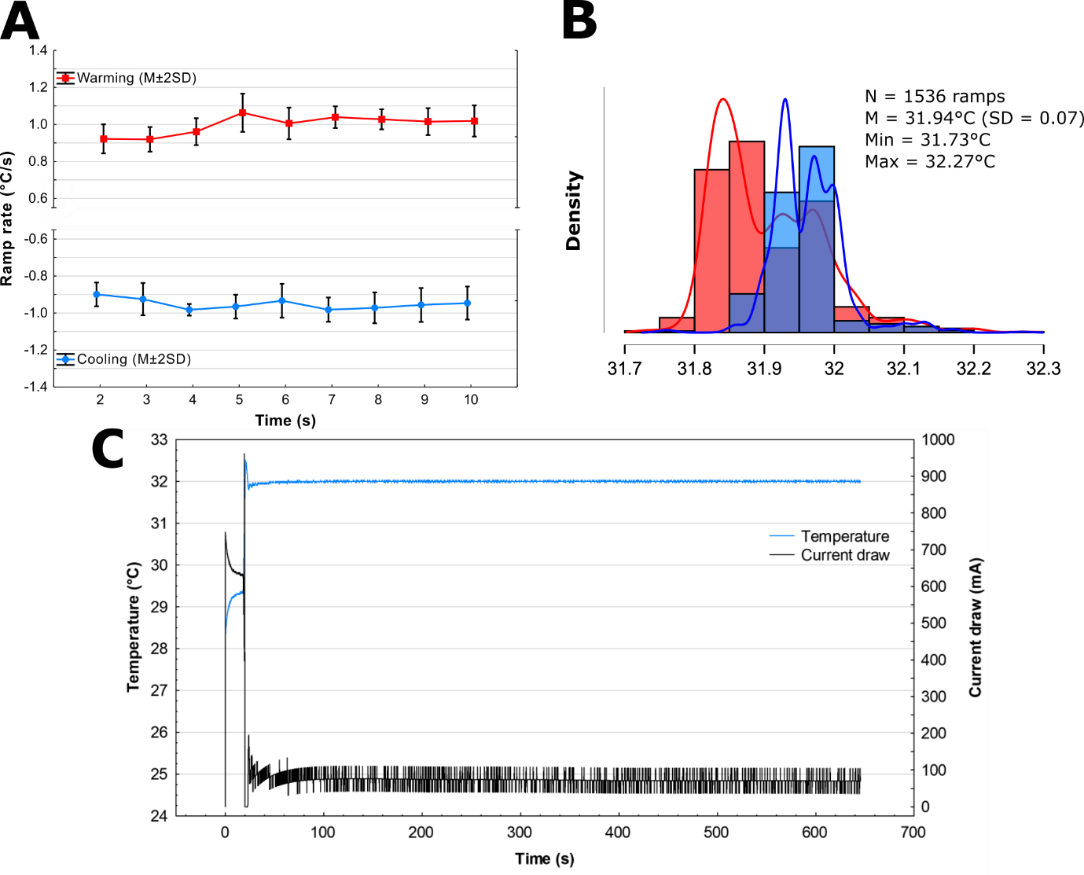

Hand placed on stimulation pad

**Fig S1**. Detailed thermal performance of SenseCheq. A – ramp rate stability plotted second-by second from 25 full-length ramps (after accounting for initial lag as seen in Fig 3). The second-by-second rates are stable around -1 and +1 °C/s for cooling and heating respectively. B – ramp starting temperatures across 1536 ramps from 192 sessions across 6 devices conducted unsupervised at home by 16 study participants for cooling (blue) and warming (red). The mean starting temperatures cluster around 31.9 °C, which is a consequence of participants’ hands typically being slightly cooler than 32 °C, the device reaching the stability threshold while in warming mode, and a gentle integral-term coefficient which was chose as a reduced overshoot was prioritized over hitting the set point as quickly as possible. C – a characteristic thermal stabilisation curve for SenseCheQ, stabilising to and maintaining 32 °C for 600 seconds.

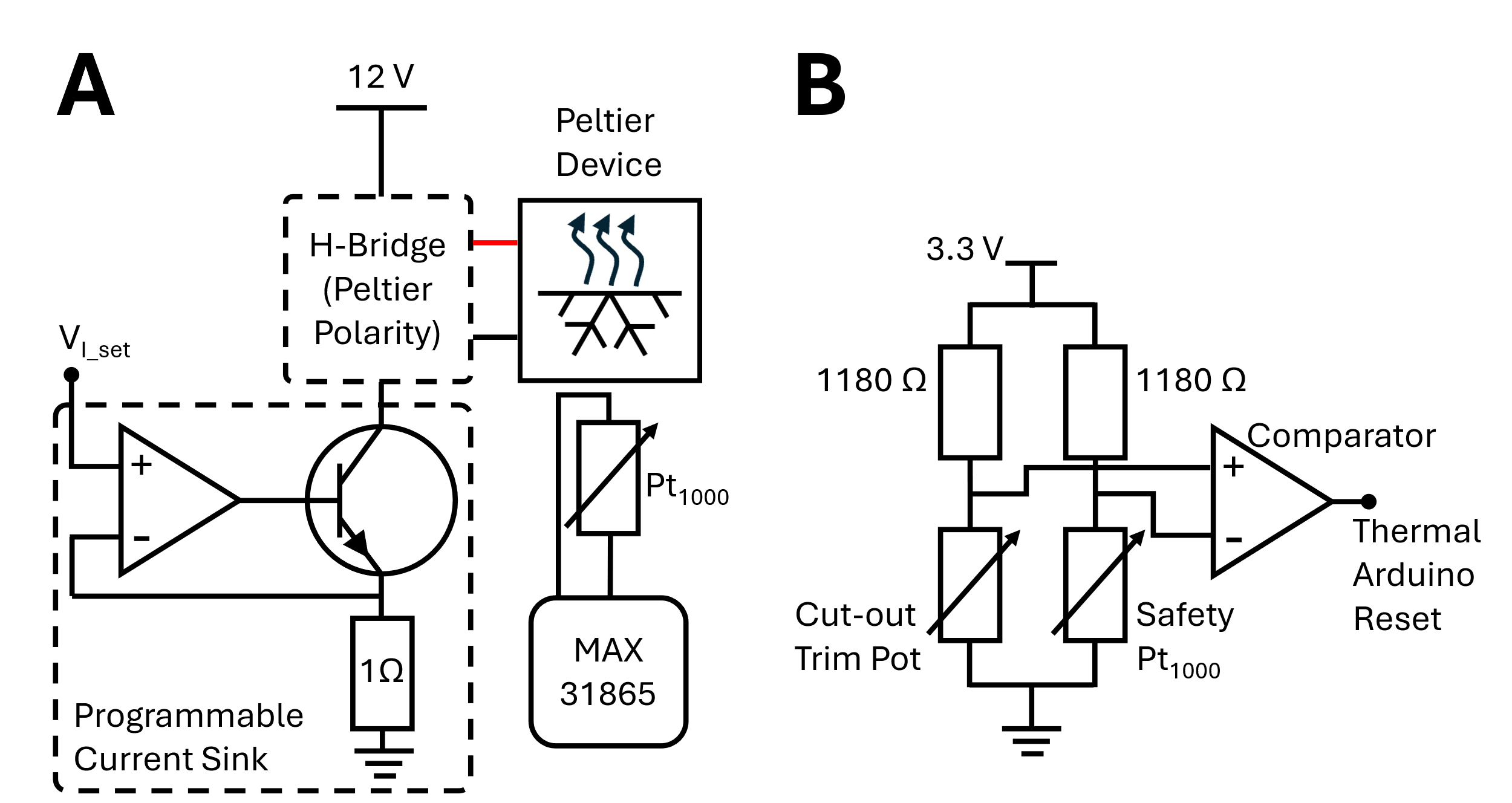

**Fig S2**. Detailed circuit diagrams for thermal control: A – the Peltier drive and control system. The Pt1000 in combination with the Pt1000 amplifier (MAX31865) provide the closed-loop feedback for the proportional-integral thermal control. The programmable current sink comprises a zero-drift op-amp (OPA189), a BDX33C Darlington Pair and a 1 Ω reference resistor, which provides a one-to-one conversion between the voltage supplied at I_set_ and the current draw through the Peltier drive circuitry. The H-Bridge allows the direction of current through the Peltier to be reversed, to allow heating and cooling of the faceplate; B – the safety cut-out comparator circuit. The Pt1000 for the safety circuit is mounted on the thermal faceplate next to the closed-loop feedback Pt1000. A comparator (AP331A) reads the two central nodes of a Wheatstone bridge, wherein the safety Pt1000 is monitored in inverting mode against a variable/trim resistor. During the construction and validation of a SenseCheQ device, the trim resistor is set such that if the resistance of the safety Pt1000 increases above 1176 Ω (equivalent to 45 °C), the comparator sets its output low. The comparator’s output is tied to the reset pin of the thermal control microcontroller, thus pulling the comparator’s output low sets the output of the thermal circuit to 0. Because a high temperature on the external side of the Peltier means that the internal side of the Peltier is cool, this quickly cools the thermal face plate to a non-noxious temperature.
